# Survey of Physician Attitudes on Artificial Intelligence

**DOI:** 10.64898/2026.11.01.26367027

**Authors:** Lisa Leininger, Tasha Glenn, Heather Mayle, Nyla Hart, Claire Muckian, Yagna Reddy, Kathryn Redinger, Scott Monteith, Joseph F. Goldberg, Daniel Tuinstra, Elaine Yeung, Paul Fu, David Gratzer, Charlotte Blease, Ilang M. Guiroy, John Torous, Patricia Marcy, John M. Kane, Eric D. Achtyes

## Abstract

**Objective:** To better understand how physicians view the potential use of artificial intelligence (AI) in their practices.

**Method:** We conducted an online cross-sectional survey of 1,258 physicians (MD, DO, MBBS or equivalent) in the United States (US) and Canada from December 2023 through February 2026. The survey contained 34 questions focused on 6 areas: (1) demographics, (2) training, (3) clinical benefits, (4) clinical concerns, (5) clinical AI prediction tools, and (6) generative AI.

**Results:** A total of 130 completed surveys were received (response rate, 10%). Top specialties included psychiatry (46.2%), internal medicine (14.6%) and pediatrics (13.8%). A total of 16.9% had training in computer science or information technology and 40.8% in statistics. Nearly half (46.9%) had no formal training in AI and were self-taught. Those with training in AI received it through vendors or at work. Sixty to seventy percent thought AI would provide diagnostic aid or treatment recommendations, but half did not feel adequately trained to safely use AI in practice. Over 70% thought it could help with administrative tasks. More than 80% were worried about errors, misinformation, or biases in the data.

**Conclusion:** There is a growing sense that AI in medicine could be helpful with specific tasks. Ongoing training, education, and study are needed to determine how AI will best integrate into clinical practice.

Artificial intelligence (AI) is rapidly being marketed for and integrated into clinical medicine. Previous physician surveys have shown that physicians are optimistic, yet cautious, about its implementation.

This study utilizes a novel survey tool to assess physician attitudes toward the integration of AI in practice, highlighting the desire for ongoing education around and monitoring of AI among physicians in the United States and Canada.

There is a growing recognition that AI, while a powerful tool, may harbor biases, promulgate misinformation, and lead to harm to patients, and requires ongoing training and oversight by humans.

## Introduction

A major achievement of the 20th century was the development of algorithms included in both the hardware and software that routinely power much of daily life.^1^ Algorithms based on artificial intelligence (AI) are now transforming diverse domains including commerce, education, logistics, transportation, and healthcare. In early 2026, it was estimated that about 70% of businesses use AI technologies, with the percentage expected to increase.^2^ AI enables computing technology to simulate human learning, problem solving, and creativity, with AI decisions being made instantly and locally.^3^ The value of AI algorithms is determined by the data used across the AI lifecycle, with the diverse dimensions of data quality including accuracy, completeness, consistency, integrity, and timeliness.^4^ AI algorithms that are used in medicine must be trained to be accurate and consistent, updated as needed to both reflect current practice guidelines and match local patient demographics, and deployed in a manner that allows productive use by varying levels of staff. The use of AI algorithms in medicine is expected to improve patient outcomes by enabling early detection, disease prediction, risk stratification, and personalized treatment strategies.^5^ Estimates of AI use for mental health vary, ranging from less than 3% of the population^6^ to over 50% among psychologists.^7^ The goal is for AI to enable individual healthcare approaches and early interventions, optimize clinical decision making, and improve ongoing patient monitoring. Physicians are also routinely exposed to commercial advertising of AI-based products.^8^ The purpose of this study was to survey physician understanding and routine use of AI in clinical practice, to better understand how physicians perceive the impact of AI on their professional life and patient care.

## Method

### Survey

An online survey to assess physicians’ attitudes toward the use of AI in medicine was developed using LimeSurvey software Version 6.3.6, which could be completed from a Windows, Mac, or smartphone device using a web browser.^9^ The survey contained 34 questions focused on 6 areas: (1) demographics, (2) training, (3) clinical benefits, (4) clinical concerns, (5) clinical AI prediction tools, and (6) generative AI. A pilot survey was first completed by several volunteer resident physicians, and questions were refined based on the feedback and results. The survey questions and complete response summary are provided in the Supplemental File.

### Recruitment

Licensed physicians across several US states and Canada with expertise in multiple specialties were invited to participate in the survey by email. Those who responded to the invitation were sent an email containing the link to the survey and assigned a unique token to complete the survey anonymously. There were no financial incentives for participating in the survey. All participation in the survey was voluntary. Data were collected between December 2023 and February 2026. Because tokenized survey links were sent to potential subjects and an honest broker sent only de-identified data to the research team, the study was judged not to constitute human subjects research by the Western Michigan University Homer Stryker M.D. School of Medicine Institutional Review Board (IRB).

### Statistics

The responses to all survey questions were analyzed using PSPP Version 1.6.2. The analysis included descriptive statistics of the study participants along with multiple analyses of the survey responses. A P value of 0.05 was considered to be statistically significant. No adjustments were made for multiple comparisons.

## Results

Of 1,258 individual invitations sent by email to physicians, there were 130 completed surveys, for a 10% response rate. Similar response rates of <12% were reported in other physician surveys, even when financial incentives were provided.^10,11^ The response rate obtained varied by physician specialty, with the largest numbers from psychiatry (n=60; 46.2%), internal medicine (n=19; 14.6%), and pediatrics (n=18; 13.8%). The demographic characteristics of the 130 physicians who completed the survey are shown in Table 1. Of the 130 physicians, 22 (16.9%) had formal training in computer science or information technology, and 53 (40.8%) had formal training in statistics. Physicians born before 1975 were more likely to have formal training in computer science or information technology than those born 1975 or after (P=0.023). There was no significant difference between males and females in formal training in technology. In this sample, 61 (46.9%) of the physicians had no training in AI and were self-taught. Psychiatrists were more likely to have received training in AI than physicians in other specialties (P=0.028). Of those who received training in AI, the sources were vendors, staff at work, and medical co-workers, with fewer than 15 physicians in each category. The ongoing sources for information about AI that physicians currently use include web browsing (64.6%), newspaper articles (55.4%), co-workers (49.2%), medical journals (48.5%), social media (45.4%), and friends or family (41.5%). Only 2.3% do not read about AI. The current level of burnout reported by the 130 physicians who completed the survey was mild, negligible, or none for 75.4% of the physicians, and moderate or severe for 24.6% of the physicians.

**Table 1.** Demographic characteristics (N=130)

| Demographic Category | N | % |
|---|---|---|
| Year of Birth |  |  |
| Born before 1975 | 60 | 48.8 |
| Born 1975 or later | 63 | 51.2 |
| Gender |  |  |
| Male | 74 | 56.9 |
| Female | 55 | 42.3 |
| Race |  |  |
| White | 97 | 74.6 |
| Black/African American | 4 | 3.1 |
| Asian | 23 | 17.7 |
| Year Graduated Medical School |  |  |
| Before 2005 | 64 | 50.0 |
| 2005 or later | 64 | 50.0 |
| Type of Practice |  |  |
| Private practice, specialty group, community health clinic, HMO | 35 | 26.9 |
| Academic, hospital based or primarily research, other | 95 | 73.1 |
| Affiliated with a Medical School |  |  |
| Full time faculty | 51 | 39.2 |
| Part time faculty, community faculty | 36 | 27.7 |
| Resident, fellow | 14 | 10.8 |
| No | 29 | 22.3 |

Overall, about 70% of all physicians expect that in the future AI will be providing diagnosis decision support, and 61.8% expect treatment recommendations. Yet, of those surveyed, about half (49.6%) felt they did not have sufficient background to safely and effectively use AI in their daily practice. Additionally, 88.5% of the physicians felt that the ongoing medical education they received did not provide sufficient training in AI, and 80% worried about false negatives or false positives when using AI prediction tools. Furthermore, 84.6% of physicians are concerned about bias or errors in the data used to train AI, specifically noting errors in EMR systems. There is clear recognition of many potential sources of errors in the training data, including coding errors, incomplete data, inconsistencies, biases, and lack of vendor interoperability. Sixty-four percent of physicians were concerned about biases in AI predictions due to a lack of diversity in the training data. Two-thirds of physicians (66.2%) were aware of potential privacy issues with the use of clinical AI prediction tools. Yet despite awareness of AI limitations, many physicians have a positive view of the potential benefits of AI in medicine. For example, 73.1% of the physicians felt AI could streamline administrative tasks, 55.4% felt AI would improve overall clinical decision making, and 48.5% felt investment in AI should keep increasing. In the future, the majority of physicians expect clinical AI to be used to generate clinical progress notes, disease screening, decision support for diagnosis, and interpretation of lab tests and imaging. About 78% of physicians expect AI to assist in generating patient progress notes in the EMR. Most physicians (82.3%) are also concerned that patients are getting medical misinformation from generative AI that has not been curated by a physician.

## Discussion

With the continued expansion of AI in medicine, there is a clear need to increase physician education about AI. For example, in an AMA survey of 1,692 physicians in 2026, 81% were using AI, up from 66% in 2024,^12^ and in a survey of 3,151 Doximity physicians, 63% reported use of AI in clinical practice, up from 47% in the spring of 2025.^13^ In the current survey, the majority of physicians feel that AI should be used to assist with administrative tasks, in agreement with the AMA and Doximity findings.^12,13^ The majority of physicians in the current survey do not use AI for clinical prediction. Before the introduction of AI systems into healthcare for clinical use such as decision support, robust and comprehensive evidence generation was required.^14^ While most physicians feel AI will decrease errors in clinical decision making, they are also aware of the methodological challenges associated with AI. These challenges include limitations and biases in the training data and in the generalizability of findings, and AI model reasoning failures.^15,16^ In some medical situations, human expertise has been found more reliable than AI, such as for triage in the emergency department.^17^ Successful implementation of AI in clinical medicine requires understanding of the limitations, biases, boundaries, and potential consequences, as well as data privacy assurances and ongoing oversight. A specific guideline and process needs to be in place for evaluating and implementing clinical decision support AI systems in real-world settings.^14^

There is agreement by the majority of physicians about the risks of the general public using online AI to obtain medical advice, which is especially of concern in urgent situations.^18,19^ Error rates of 20% have been found in studies of publicly available AI models that answer health-related questions.^20^ Studies of the use of AI triage systems by the general public have found missed high-risk emergency and crisis situations.^19^ Yet physicians should expect that patients will look online for medical information. There needs to be a process and regulatory framework in place to protect the general public from incorrect and inappropriate online AI medical advice, such as may come from chatbots.^21^

## Limitations

The survey participants may not be representative, as high-performing physicians are more likely to participate in research studies.^22,23^ The potential for response bias, such that the answers given did not accurately represent responder experiences, was not discussed.^24^ The potential for non-response bias, such that there were differences between those who did and did not respond to the survey, was also not discussed.^24^ Some physicians may be unaware of the limitations of AI in medicine beyond training data issues, such as how changes in the standard of care impact recommendations based on older data.^15^ The impacts of the increasing use of AI on hardware models, and the growth of domain-specific AI models, were not discussed. The increase in AI research in industry rather than academia was omitted.^25^ The majority of the physicians are aware that there are data privacy issues related to AI, although measures to address privacy issues were not included in the study. The biases generated by AI were not discussed, such as AI models of hospital administration overrepresenting males and underrepresenting diversity.^26^ It took just over two years to collect the data, and physician opinions and use of AI can shift quickly over time.^27,28^

## Conclusion

Most physicians view AI technologies as a potential approach to improve patient care, enabling disease detection and clinical decision making. Most physicians expect growing adoption of AI across medicine and predict this will improve patient care and physician productivity. Increased physician training in the use and impacts of AI technologies in medicine is needed.

## Supporting information

Physician AI survey

## Data Availability

All data produced in the present study are available upon reasonable request to the authors.

## Previous Presentation

None.

## Acknowledgments

None.

## References

1. Jordan M. A collectivist, economic perspective on AI. arXiv. Preprint posted online 2025. doi:10.48550/arXiv.2507.06268

2. Yotzov I, Barrero JM, Bloom N, et al. Firm Data on AI. National Bureau of Economic Research; 2026 Feb 16.

3. Stryker C, Kavlakoglu E. What is AI? IBM. 2026. https://www.ibm.com/think/topics/artificial-intelligence

4. Aquino J, Jonker A. Why AI data quality is key to AI success. IBM. 2026. https://www.ibm.com/think/topics/ai-data-quality

5. Ng A, Hinton G, Jordan MI. The role of artificial intelligence in personalized and precision medicine: transforming one-size-fits-all treatment into individualized care. 2026.

6. How people use Claude for support, advice, and companionship. Published online July 27, 2025. https://www.anthropic.com/news/how-people-use-claude-for-support-advice-and-companionship.

7. American Psychological Association. AI in the therapist’s office: 2025 Practitioner Pulse Survey. American Psychological Association; 2025. https://www.apa.org/pubs/reports/practitioner/2025

8. Monteith S, Glenn T, Geddes JR, Whybrow PC, Achtyes ED, Bauer R, Bauer M. Patient and physician exposure to artificial intelligence hype. Pharmacopsychiatry. Published online 2025 May 12.

9. LimeSurvey. 2026. https://www.limesurvey.org/

10. Hawa S, Bane S, Kinsler K, Rector A, Chaichian Y, Falasinnu T, Simard JF. Impact of incentives on physician participation in research surveys: randomized experiment. JMIR Form Res. 2024 May 14;8:e54343.

11. Cook DA, Wittich CM, Daniels WL, West CP, Harris AM, Beebe TJ. Incentive and reminder strategies to improve response rate for internet-based physician surveys: a randomized experiment. J Med Internet Res. 2016 Sep 16;18(9):e244.

12. AMA Center for Digital Health and AI. 2026 Physician Survey on Augmented Intelligence. American Medical Association; 2026 Mar. https://www.ama-assn.org/system/files/physician-ai-sentiment-report.pdf

13. Doximity. State of AI in Medicine. 2026 Jan. https://www.doximity.com/reports/state-of-ai-medicine-report/2026

14. Vasey B, Nagendran M, Campbell B, et al. Reporting guideline for the early stage clinical evaluation of decision support systems driven by artificial intelligence: DECIDE-AI. BMJ. 2022 May 18;377:e070904.

15. Monteith S, Glenn T, Geddes JR, Whybrow PC, Achtyes E, Bauer R, Bauer M. Be aware of AI limitations. Pharmacopsychiatry. Published online 2026 Jan 9.

16. Song P, Han P, Goodman N. Large language model reasoning failures. arXiv. Preprint posted online 2026 Feb 5. arXiv:2602.06176.

17. Zaboli A, Brigo F, Brigiari G, Massar M, Parodi M, Pfeifer N, Magnarelli G, Turcato G. Chat-GPT in triage: still far from surpassing human expertise—an observational study. Am J Emerg Med. 2025 Jun 1;92:165–171.

18. Bean AM, Payne RE, Parsons G, et al. Reliability of LLMs as medical assistants for the general public: a randomized preregistered study. Nat Med. Published online 2026 Feb 9:1–7.

19. Ramaswamy A, Tyagi A, Hugo H, et al. ChatGPT Health performance in a structured test of triage recommendations. Nat Med. Published online 2026 Feb 23:1–1.

20. Mingole B, Majumdar A, Choudhury FA, Kraschnewski JL, Sundar SS, Yadav A. Dr GPT will see you now, but should it? Exploring the benefits and harms of large language models in medical diagnosis using crowdsourced clinical cases. arXiv. Preprint posted online 2025 Jun 13. arXiv:2506.13805.

21. Chow JC, Li K. Large language models in medical chatbots: opportunities, challenges, and the need to address AI risks. Information. 2025 Jun 27;16(7):549.

22. Elston DM. Participation bias, self-selection bias, and response bias. J Am Acad Dermatol. Published online 2021 Jun 18.

23. Dahrouge S, Armstrong CD, Hogg W, Singh J, Liddy C. High-performing physicians are more likely to participate in a research study: findings from a quality improvement study. BMC Med Res Methodol. 2019 Aug 7;19(1):171.

24. Sedgwick P. Non-response bias versus response bias. BMJ. 2014 Apr 9;348:g2573.

25. Eastwood J. Study: industry now dominates AI research. MIT Sloan. 2023. https://mitsloan.mit.edu/ideas-made-to-matter/study-industry-now-dominates-ai-research

26. Gisselbaek M, Berger-Estilita J, Minsart L, et al. Gender disparities in artificial intelligence– generated images of hospital leadership in the United States. Mayo Clin Proc Digit Health. 2025 Jun 1;3(2):100218.

27. Blease C, Hagström J, Sanchez CG, et al. General practitioners’ adoption of generative artificial intelligence in clinical practice in the UK: an updated online survey. Digit Health. 2025 Nov;11:20552076251394287.

28. Blease CR, Locher C, Gaab J, Hägglund M, Mandl KD. Generative artificial intelligence in primary care: an online survey of UK general practitioners. BMJ Health Care Inform. 2024 Aug 29;31(1):e101102.

