## Supplementary material for "Survey of Physician Attitudes on Artificial Intelligence": Physician AI survey

### DEMOGRAPHICS

What is your race? (Check all that apply)

- White
- Black/African American
- American Indian
- Alaska Native
- Native Hawaiian
- Other Pacific Islander
- Asian
- Don't know

What is your medical specialty? (Check all that apply)

- Anesthesiology
- Dermatology
- Family practice/general medicine
- Internal medicine
- Medical Genetics
- Neurology
- Obstetrics & Gynecology
- Pathology
- Pediatrics
- Psychiatry
- Radiology
- Rheumatology
- Surgery

What best describes the type of practice are you in? (Check one)

- Solo (private practice)
- Single specialty group
- Multi-specialty group
- Health Maintenance Organization (HMO) group
- Community Health Clinic
- Academic
- Hospital-based
- Primarily research
- Other

Are you currently affiliated with a medical school? (Check one)

- Yes, full time faculty appointment
- Yes, part time faculty appointment
- Yes, community faculty appointment
- Yes, fellow/resident
- No

Of the time spent providing patient care in a typical week, what percent of time do you spend completing required administrative tasks? (Check one)

- < 25%
- 25-50%
- 50-75%
- > 75%

Approximately how many hours a week do you typically spend on patient care, including the required administrative duties? (Check one)

- 0-15
- 16-30
- 31-45
- 46-60
- > 60

Many recent articles have reported widespread physician burnout. How would you rate your current level of burnout? (Check one)

- Severe
- Moderate
- Mild
- Negligible
- None

Sex at birth? (Check one)

- Male
- Female
- Prefer not to answer

Is your ethnicity Hispanic, Latino/a, or Spanish? (Check all that apply)

- Mexican, Mexican American, Chicano/a
- Puerto Rican
- Cuban
- Another Hispanic, Latino/a, or Spanish ethnicity
- No
- Don't know

TRAINING

Have you had any formal training in any of these areas? (Check all that apply)

- Computer science or information technology
- Data science
- Statistics
- Operations research
- None of the above

What type of training have you had in artificial intelligence (AI)? (Check all that apply)

- College courses
- Self-taught (No formal training)
- Vendor provided training
- Training provided by technology staff at work
- Training provided by medical co-workers
- External training courses
- None

What are your ongoing sources of information about AI? (Check all that apply)

- Newspaper articles
- Online help and user guides
- Social media
- Medical journals
- Technical journals
- Vendors
- Online media/web browsing
- Co-workers
- Friends or family
- Don't read about AI

Do you think you have sufficient background in AI to safely and effectively use AI tools in your daily practice? (Check one)

- Yes
- No
- Not sure

In your experience, has continuing medical education provided sufficient training in AI? (Check one)

- Yes
- No
- Not sure

When would you trust learning about AI tools from a vendor promoting the tool? (Check one)

- Only if the clinical AI prediction tool had regulatory approval with proof of safety and efficacy
- With endorsement from my professional medical society
- Never - worried about vendor conflict of interest
- Not sure

CLINICAL BENEFITS

Which of these statements about how clinical AI prediction tools will improve medicine in the future do you agree with? (Check all that apply)

- Decrease medical errors
- Increase access to screening
- Improve overall quality of care
- Improve disease outcomes
- Improve disease outcome predictions
- Relieve physician administrative burdens
- Decrease reliance on specialists
- None of the above
- Don't know

Which of the following tasks do you expect clinical AI prediction tools to perform or assist in performing in medicine in the future? (Check all that apply)

- Screening for disease
- Risk stratification
- Diagnosis decision support
- Treatment recommendations
- Prediction of disease outcomes
- Generation of patient progress notes in the electronic medical record (EMR)
- Lab test and/or digital imaging interpretation
- Remote patient monitoring
- Genetic analyses
- None of the above
- Don't know

Do you think that clinical AI prediction tools will help extend care to remote and/or under served areas in the future? (Check one)

- Yes
- On a limited basis
- No
- Don't know

Which of these statements about the overall role of AI in medicine do you agree with? (Check all that apply)

- AI will revolutionize clinical medicine
- AI will expand healthcare services
- AI will improve overall clinical decision making
- AI will improve my clinical decision making
- AI will streamline administrative tasks
- Investment in AI in medicine should keep increasing
- None of the above
- Don't know

CLINICAL CONCERNS

Which of the following problems are you concerned about with the use of clinical AI prediction tools? (Check all that apply)

- False negative or false positive predictions
- Unknown safety and efficacy if not Food and Drug Administration (FDA) approved
- Bias or errors in the training data
- Lack of education or training in AI
- Privacy concerns
- Uncertain legal liability for AI errors
- Cybersecurity risks
- None of the above
- Not sure

Which of the following limitations of EMR data are you concerned will affect training of clinical AI prediction tools? (Check all that apply)

- Inaccurate or incomplete data
- Coding errors in the EMR
- Biases due to a lack of patient diversity including age, sex, race, illness severity
- Interoperability issues between vendor EMR products
- Changing treatment recommendations over time
- Geographic or socioeconomic differences in treatment
- Inconsistent patient data (lab tests, imaging, remote monitoring devices/apps)
- None of the above
- Not sure

Which of the following problems are you concerned about when data from patient digital apps are used to train clinical AI prediction tools in medicine? (Check all that apply)

- Electronic sensor properties lead to measurement inaccuracies and inconsistencies
- Different manufacturers use different electronic sensors
- Electronic sensors differ between makes and models from the same manufacturer
- Patient behaviors contribute to measurement errors
- Patients may not use apps as directed
- Data collected by different software versions
- Data collected by different software products
- Patients change smartphone manufacturers or models
- None of the above
- Don't know

#### CLINICAL AI PREDICTION TOOLS

What do you think about your use of clinical AI prediction tools in medicine? (Check all that apply)

- AI will help me with the diagnosis and treatment of difficult cases

AI will ease my administrative burden
I am not sufficiently knowledgeable to use AI safely
I am worried about losing my clinical skills
I am concerned about becoming dependent on technology
None of the above
Don't know

To your knowledge, are you currently using any clinical AI prediction tools? (Check one)
Yes
No
Don't know

If yes, who developed the clinical AI prediction tool (Check one)
Part of EMR
Custom built tool
Vendor outside of EMR
Don't know

After an examination of a patient, a clinical AI prediction tool provided a specific diagnosis that was different from your diagnosis. How would you react? (Check one)
Accept the AI diagnosis as correct
Accept the AI diagnosis if the tool was FDA approved, or validated by my specialty organization, or another regulatory agency
Consider the AI diagnosis, but stay with your diagnosis based on the patient examination
Consider the AI diagnosis, and re-evaluate the patient
Ignore the AI diagnosis
Not sure

Are you concerned that the increasing use of clinical AI prediction tools will decrease your diagnostic and/or physical examination clinical skills? (Check one)
Yes
No
Not sure

Are you concerned that you will develop over reliance on clinical AI prediction tools? (Check one)
Yes
No
Not sure

Does it concern you that most clinical AI prediction tools do not have regulatory approval, including proof of safety and efficacy? (Check one)
Yes
No
Not sure

Does your practice document and track errors associated with clinical AI prediction tools? (Check one)
Yes
No
Not sure
Not applicable

GENERATIVE AI

Do you use or plan to use generative AI models such

as ChatGPT? (Check all that apply)

- For fun and tasks not related to work
- Tried just to gain understanding of these models
- Do not use or plan to use

Which areas of healthcare do you think generative AI models such as ChatGPT will be routinely used in the future? (Check all that apply)

- Patient appointment scheduling
- Patient symptom checkers
- Create insurance company appeals and claims processing
- Create patient notes
- Provide answers to physicians on complex medical questions
- Create summaries of recorded patient physician conversations
- Remote patient monitoring
- Help writing scholarly publications
- Assist with teaching/presentations for learners
- None of the above
- Don't know

Do you agree with any of the following concerns with generative AI? (Check all that apply)

- Generative AI output often contains misinformation
- Misinformation often appears correct to those who are not expert in the field
- Fabricated references are often created for misinformation
- Misinformation appears customized for the individual patient
- Generative AI models are easy for patients to use
- Patients do not consult with a medical professional
- Patients use AI to disagree with providers
- Patient misuse of medical resources based on misinformation
- None of the above

Are you concerned that patients are getting medical misinformation from generative AI models that has not been curated by a medical professional? (Check one)

- Yes
- No
- Not sure
